# Estimation of Daily Reproduction rates in COVID-19 Outbreak

**DOI:** 10.1101/2020.12.30.20249010

**Authors:** Jacques Demongeot, Kayode Oshinubi, Hervé Seligmann, Florence Thuderoz

**Affiliations:** Laboratory AGEIS EA 7407, Team Tools for e-Gnosis Medical and Labcom CNRS/UGA/OrangeLabs Telecom4Health, Faculty of Medicine, Université Grenoble Alpes (UGA), 38700 La Tronche, France; The National Natural History Collections, The Hebrew University of Jerusalem, 91404 Jerusalem, Israel; Institute of Microstructure Technology, Karlsruhe Institute of Technology (KIT) Hermann-von-Helmholtz-Platz 1, 76344, Eggenstein-Leopoldshafen, Germany

**Keywords:** daily reproduction rate, Covid-19 outbreak, discrete epidemic growth equation, discrete deconvolution

## Abstract

**(1) Background:** The estimation of daily reproduction rates throughout the infectivity period is rarely considered and only their sum *R*_*o*_ is calculated to quantify the level of virulence of an infectious agent;

**(2) Methods:** We give the equation of the discrete dynamics of epidemic growth and we obtain an estimation of the daily reproduction rates, by using a technique of deconvolution of the series of observed new cases of Covid-19;

**(3) Results:** We give both simulation results as well as estimations for several countries for the Covid-19 outbreak;

**(4) Conclusions:** We discuss the role of the noise on the precision of the estimation and we open on perspectives of forecasting methods to predict the distribution of daily reproduction rates along the infectivity period.

## 1 Introduction

After the Severe Acute Respiratory Syndrome outbreak by coronavirus SARS CoV in 2002 and the Middle East Respiratory Syndrome outbreak by coronavirus MERS CoV in 2012, the Covid-19 disease by coronavirus SARS CoV-2 is the third coronavirus outbreak in the past two decades. Human coronaviruses including 229E, OC43, NL63, and HKU1 are a group of viruses that cause a significant percentage of all common colds in humans. The SARS CoV-2 can be transmitted from person to person by respiratory droplets and through contact and fomites. Therefore, the severity of disease symptoms such as cough, sputum, and their viral load are most often important factors in the virus’s ability to spread, and these factors can change rapidly in just a few days during the period of infectivity of an individual. This ability to spread is quantified by the classic basic reproduction number *R*_*o*_ (also called basic reproduction ratio or rate), an epidemiology parameter which describes the transmissibility of an infectious agent and is equal to the mean number of susceptible individuals, an infected individual can contaminate during his infectivity period. *R*_*o*_ is not a biological constant for a pathogen: in fact, *R*_*o*_ is affected by numerous exogenous factors like geoclimate, demographic and socio-behavioural factors that govern pathogen transmission [1,2,3,4,5,6].

Because of these environmental factors, *R*_*o*_ might change seasonally, but these factors variations are not significant if a very short time in considered. *R*_*o*_ depends also on endogenous factors like the viral load of the infected individuals during their infectivity period; the variations of this viral load [8,9,10,11,12,13,14] is often neglected in the theoretical and applied studies on the Covid-19 outbreak, in which the authors estimate a unique reproduction number *R*_*o*_ linked to the Malthusian growth parameter of the exponential phase of the epidemic during which *R*_*o*_ is greater than 1 (Figure 1) and they rarely consider the distribution of partial daily reproduction numbers at day *j* of the infectivity period, denoted 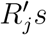 [1]. When this distribution is considered, it is more convenient i) to define for each age class the distribution of the marginal daily reproduction numbers, ii) to estimate in each case its entropy and simulate the dynamics either using a Markovian model like that defined in Delbrück’s approach [2] or an ODE SIR model. In the Markovian case, *R*_*o*_ has to be replaced by the evolutionary entropy defined by L. Demetrius as the Kolmogorov-Sinaï entropy of the corresponding random process, which has properties concerning the stability of its Markovian invariant measure, analogue to the properties of the Malthusian parameter for the stability of the ODE’s steady state [3,4]. We compare these two modelling approaches in Section 2, then present in Section 3 as illustration results of estimation of daily reproduction rates in different countries at different periods. In Section 4, we discuss our method in the framework of the classical population dynamics (Leslie model) and eventually, in Section 5, we conclude and open the approach to the multi-age model.

**Figure 1.**
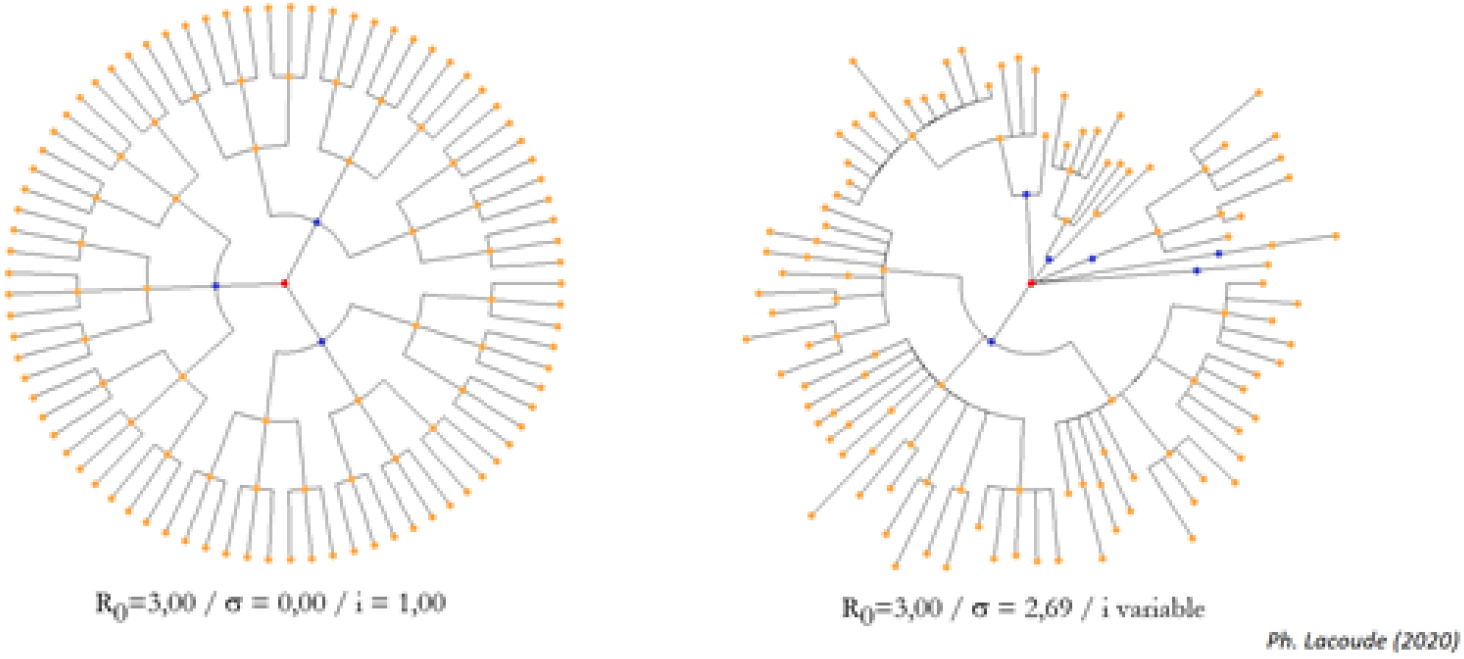
Spread of an epidemic disease from a first infected “patient zero”(in red) located on the centre of his influence sphere made of the successive generations of infected, for the same value of the reproduction number *R*_*o*_ = 3, with a deterministic dynamics (left) and a stochastic one (right) with standard deviation *σ* of the uniform distribution on an interval centred on *R*_*o*_ and with a random time interval *i* between infected generations (after [7]).

## 2 Materials and Methods

### 2.1 Relationship between Markovian and ODE SIR approach

#### 2.1.1 Obtaining SIR equation from a discrete mechanism

Let consider the classical reproduction rate *R*_*o*_. If the model is deterministic, if *X*_*j*_ denotes the number of new cases at day *j*, and the contagious period is made of *r* consecutive days, with *R*_*k*_ the marginal reproduction rate at day *k* of the contagious period, we have:

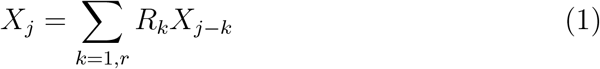

It is easy to show that, if *X*_*o*_ = 1 and *r* = 5 (inside the estimated interval of the duration of the maximal contagion period for the COVID-19 [5,6,7,8,9,10,11,12,13,14,15,16,17], we obtain:

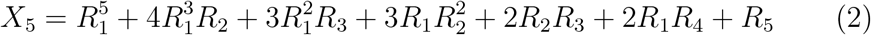

If *R*_2_ and *R*_3_ are dominant and equal to *R/*2, then *X*_5_ behaves as 2*R*_2_*R*_3_ = *R*^2^/2, which shows the difficulty to estimate *R*_*o*_, which is the mean value of the 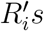 on the contagious period. The length of this period can be estimated from the ARIMA series of the stationary random variables 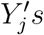, equal to the 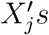 without their trend: one can take the interval on which the auto-correlation function remains more than a certain threshold, e.g., 0.1 [15]. More generally if *R*_1_ = *a, R*_2_ = *b* and *R*_3_ = *c*, we get:

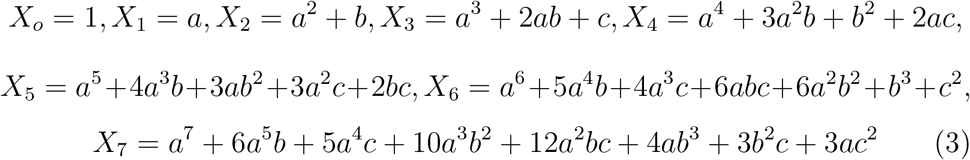

If *R*_1_ and *R*_2_ equal respectively *a* and *b*, and if *a* = *b* = *R/*2, *c* = 0, then *X*_5_ behaves like:

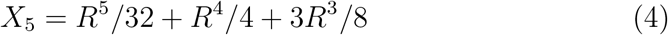

If *R* = 1, {*X*_*j*_}_*j*=1,*∞*_ is the Fibonacci sequence, and more generally, for *R >* 1, the generalised Fibonacci sequence. Let suppose now that *b* = 0 and *a* depends on the day *j* such as *a*_*j*_ = *νC*(*j*), where *C*(*j*) represents the number of possible susceptible individuals, which can be recruitable by one infectious individual at day *j*. If cumulated infected individuals (supposed to be all infectious) at day *j* are denoted by *I*_*j*_, we have:

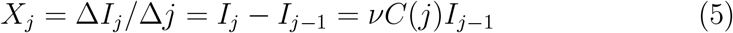

Suppose that the first infected is recruited at the centre of his sphere of influence and that the secondary infected individuals remain in this sphere, by widening the radius on day *j*, therefore the susceptible individuals *C*(*j*), where each infectious on day *j* − 1 can recruit are located on a decreasing part of the sphere of influence of the first infected 0 (Figure 2).

**Figure 2.**
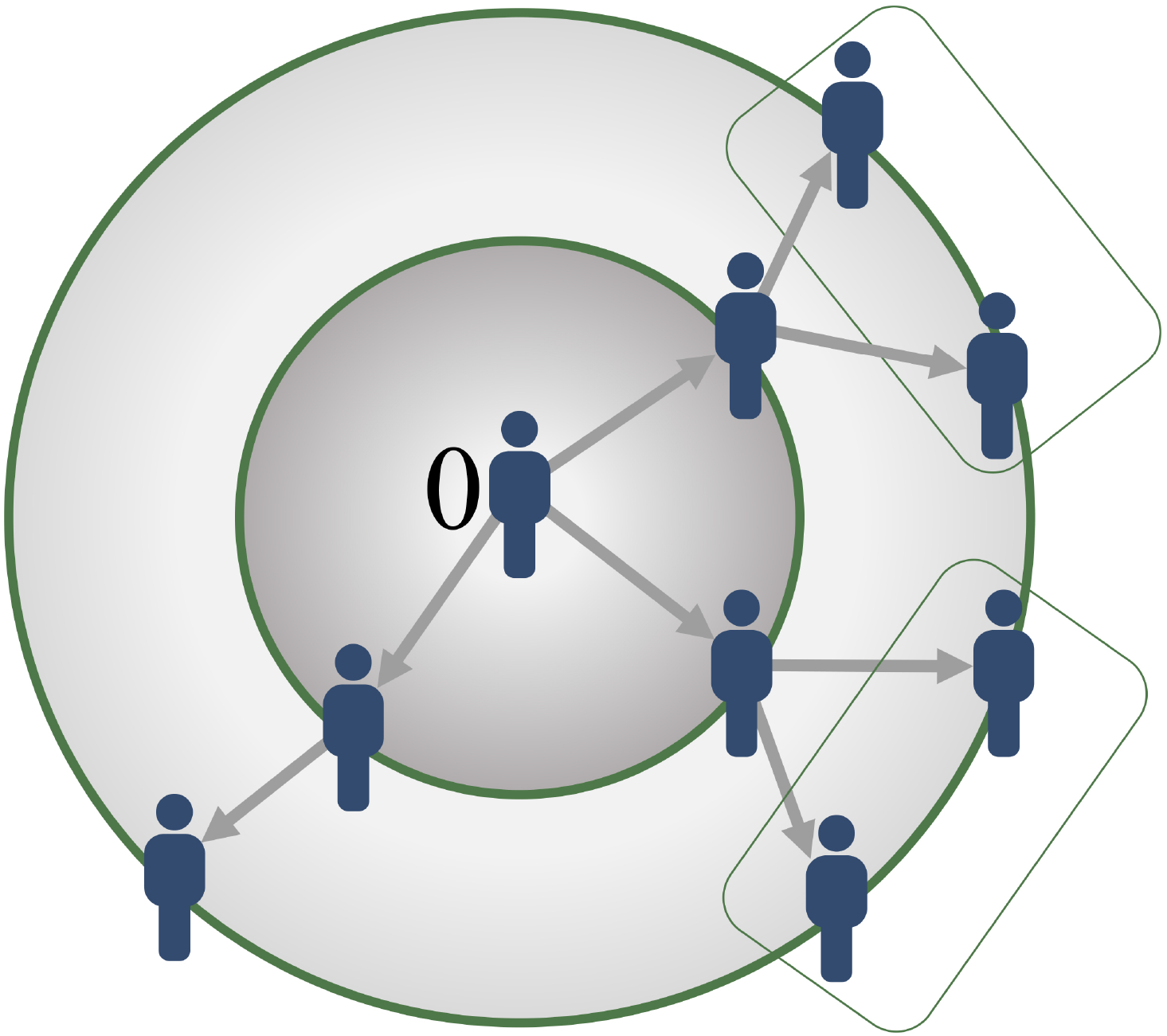
Spread of an epidemic disease from a first infected (located at his influence circle centre) progressively infecting all the neighbours in some regions (rectangles) on successive spatial layers.

The function *C*(*j*) decreases due to the bulk on the successive spheres and we can consider the following functional form *C*(*j*) = *S*(*j*)/(*c* + *S*(*j*)), where *S*(*j*) is the number of susceptible individuals at day *j*. Then, we can write the following equation taking into account the mortality:

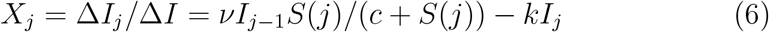

The corresponding continuous equation is close to the SIR equation, if *c* is greater before *S*:

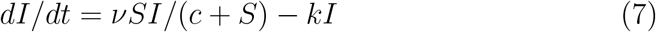

#### 2.1.2 Second obtention of the SIR equation from a discrete mechanism

Another way to derive the SIR equation from a probabilistic approach is to start from the microscopic equation of molecular shocks by Delbrück [2] which corresponds to a classical birth-and-death process: if at least one event (contact *ν*, birth *f*, death *μ* or recovering *ρ*) occurs in (*t, t* + *dt*), we have, if births compensate deaths, leaving constant the total size *N* of the population:

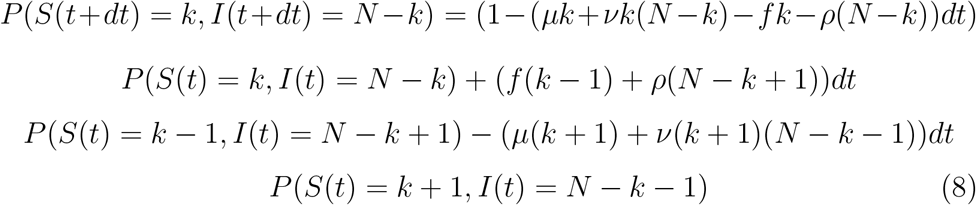

Hence, we have, if *P*_*k*_(*t*) denotes *P* (*S*(*t*) = *k, I*(*t*) = *N* − *k*):

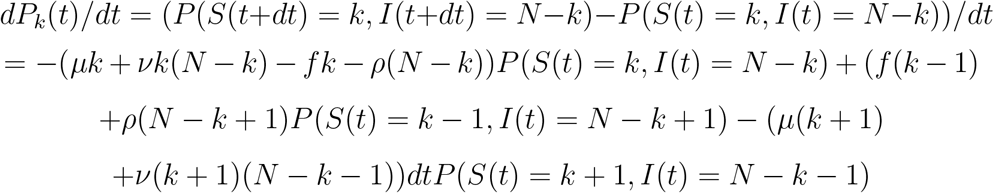

and we obtain:

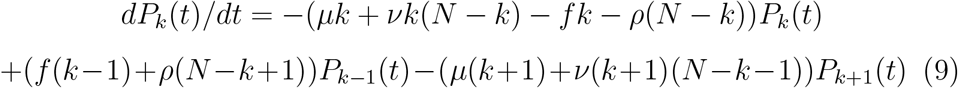

Then, by multiplying by *s*^*k*^ and summing over *k*, we obtain the characteristic function of the random variable *S*, which is proven to be a Poisson random variable if the coefficients *ν, f, μ* and *ρ* are sufficiently small. If births do not compensate deaths, we have:

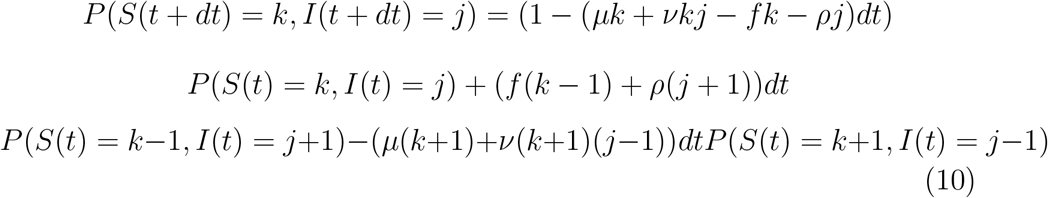

If *S* and *I* are independent and if the coefficients *ν, f, μ* and *ρ* are sufficiently small, they are Poisson random variables, whose expectations *E*(*S*) and *E*(*I*) verify:

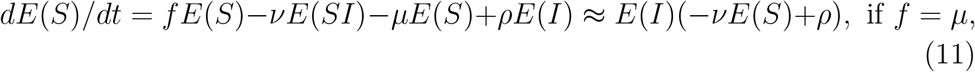

which leads to the SIR equations for the variables *S, I* and *R* considered as being deterministic:

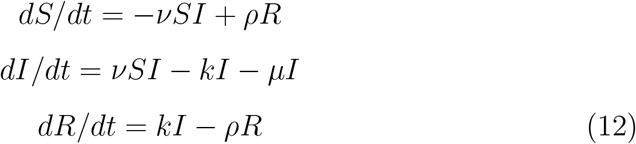

### 2.2 Distribution of the rate of transmission along the infectiousness period of an individual

Figure 3 gives the average transmission rate *R*_*o*_ calculated on the 25th of October 2020 just before the second lockdown (re-confinement) in France [16], but because at this day the second wave of the epidemic is still in its exponential phase, it is more convenient i) to consider the distribution of the marginal daily reproduction numbers, and ii) to calculate its entropy and simulate the epidemic dynamics using a Markovian model [2]. If *R*_*o*_ denotes the average transmission rate (or mean reproduction number) among the studied population, we can estimate the distribution *V* (whose coefficients are *V*_*j*_ = *R*_*j*_*/R*_*o*_) of the daily reproduction numbers *R*_*j*_ along the infectiousness period, by remarking that the number *X*_*j*_ of new infectious cases at day *j*, equal to *X*_*j*_ = *I*(*j*)–*I*(*j* − 1), where *I*(*j*) is the number of infectious at day *j*, verifies the discrete convolution equation:

**Figure 3.**
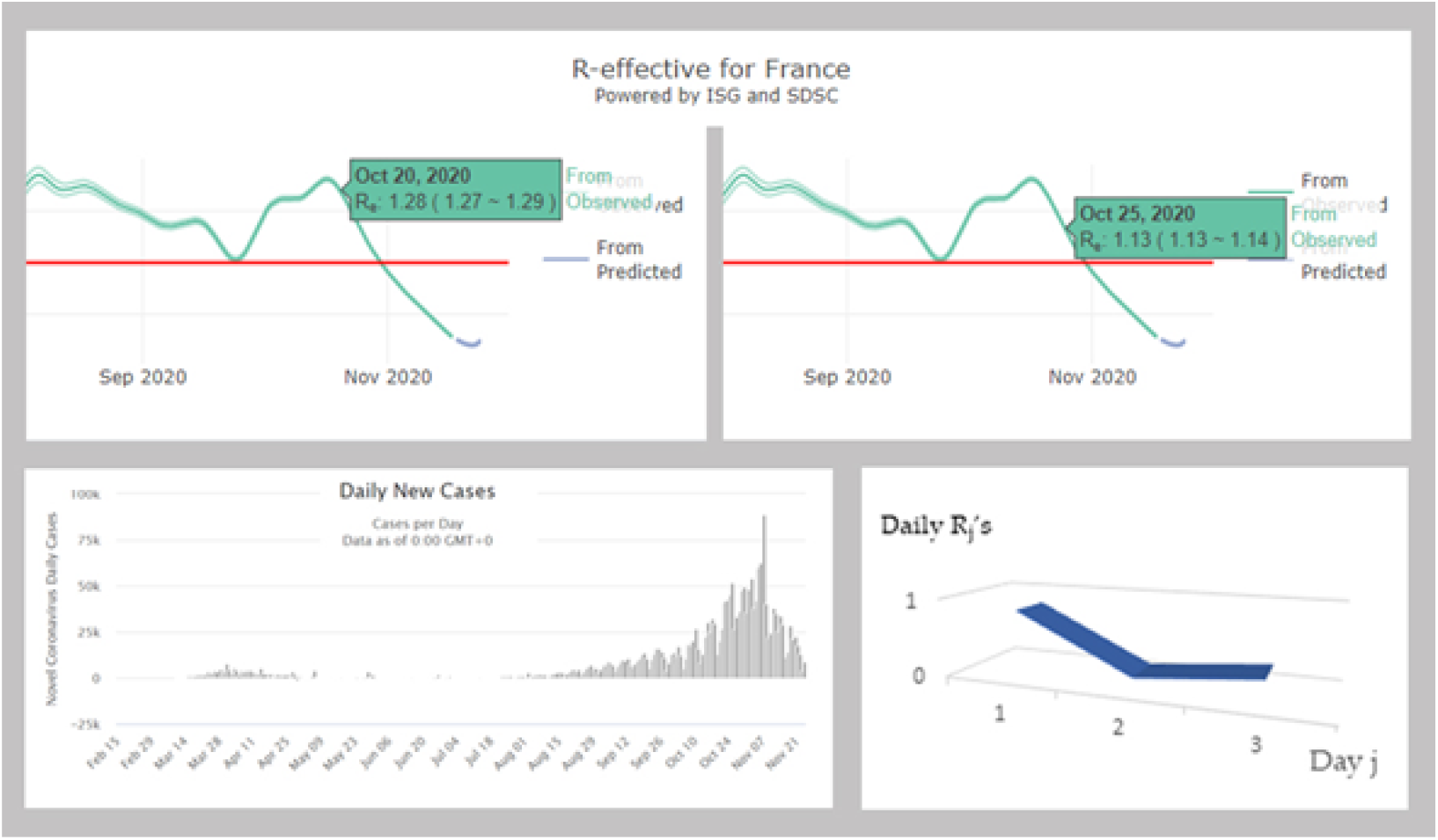
Top: estimation of the average transmission rate *R*_*o*_ for the 20th and the 25th October 2020 [18]. Bottom left: daily new cases in France between February 15 and Oct 27 [6]. Bottom right: V-shape of the evolution of the daily 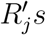 along the infectious 3-day period of an individual.

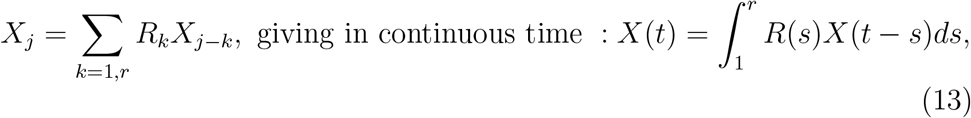

where *r* is the duration of the contagion period, estimated by 1/(*k* +*μ*), where *k* is the recovering rate and *μ* the death rate in SIR equations:

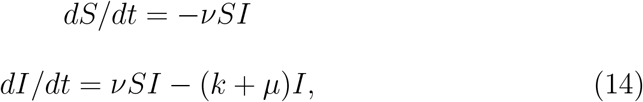

where *S* and *I* are respectively the size of the susceptible and infectious populations.

If *r* and *S* can be considered as constant during the first exponential phase of the pandemic, we can also assume that the distribution *V* is constant and then, *V* can be estimated by solving the linear system:

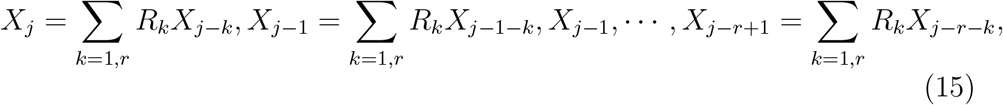

which can be written as *X* = *MR*, hence giving:

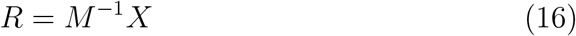

and the equation (13) can be solved numerically, if the pandemic is observed during a time greater than 1/(*k* + *μ*). Then, the entropy of *V* = *R/R*_*o*_ can be calculated, as the Kolmogorov-Sinaï entropy of the Markovian Delbrück scheme [2] ruling the 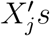 and giving new parameters for characterising pandemic dynamics, namely for quantifying its robustness and stability [3,4].

### 2.3 The biphasic pattern of the virulence curve in coronaviruses

Mostly, the clinical course of patients with seasonal influenza shows a biphasic occurrence of symptoms with two distinct peaks. Patients have a classic influenza disease followed by an improvement period and a recurrence of the symptoms [1,2,3]. The influenza RNA virus shedding (the time during which a person might be infectious to another person) increases sharply one-half to one day after infection, peaks on day 2 and persists for an average total duration of 4.5 days, between 3 and 6 days, which explains that we will choose in the following as infectivity duration these extreme values, i.e., either 3 or 6 days depending on the positivity of the estimated daily reproduction rates. It is common to see this biphasic influenza clinically: after incubation of one day, there is a high fever, then a drop in temperature before rising, hence the term “*V*” fever. The other symptoms like coughing often also have this improvement on the second day of the flu attack: after a first feverish rise (39 − 40°*C*), the temperature drops to 38°*C* on 2nd day, then rises before disappearing on the 5th day, the fever being accompanied by respiratory signs (coughing, sneezing, clear rhinorrhea, etc.). By looking the shape of virulence curves observed in coronaviruses patients [5,6,7,8,9,10,11,12], we often see this biphasic pattern.

## 3 Results

### 3.1 Distribution of the daily reproduction numbers R_*j*_’s along the infectiousness period of an individual. A theoretical deterministic example

#### Deterministic Toy Model

Let’s suppose that *a priori* 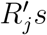 are equal to:

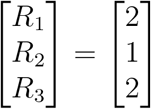

then the inverse of the matrix *M* is given by:

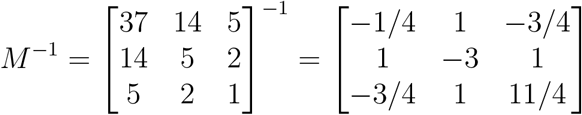

and the deconvolution gives the *a posteriori* 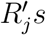:

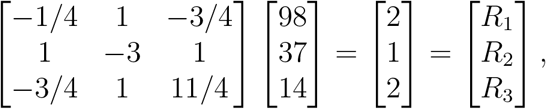

thanks to the following calculation:

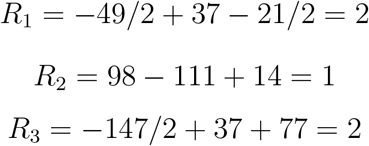

and we obtain for the *a posteriori* distribution of the daily reproduction rates the exact replica of the *a priori* distribution.

### 3.2 Distribution of the daily reproduction numbers R_*j*_’s. A simulated stochastic example

Let’s consider a stochastic version of the deterministic toy model given in Section 3.1., by introducing an increasing noise on the 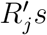, e.g., a uniform distribution on the three following intervals:[2−*a*, 2+*a*], [1−*a/*2, 1+*a/*2], [2− *a*, 2 + *a*], with increasing values of *a*, from 0.1 to 1, in order to see when the deconvolution would give negative *a posteriori* 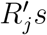, with conservation of the average of their sum *R*_*o*_, if you repeat the random choice of the values of the 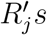 at each generation?

We will give the results of the random simulations for increasing values of *a* from *a* = 0.1 to *a* = 1.

1. **For a= 0.1**, let’s choose the *a priori* distribution of the daily reproduction numbers *R*_1_ in the interval [1.9, 2.1], *R*_2_ in [0.95, 1.05] and *R*_3_ in [1.9, 2.1] as *R*_1_ = 2.1, *R*_2_ = 0.95, *R*_3_ = 2.1. Then, transition matrix *M*_1_ is equal to:

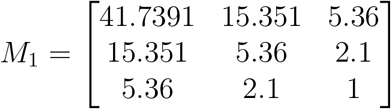

and we have:

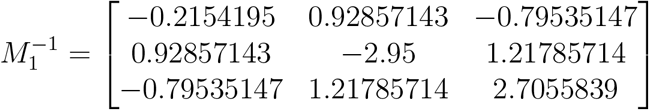 From *X*_6_ = 113.491, *X*_5_ = 41.7391, *X*_4_ = 15.351, *a posteriori* 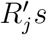 can be calculated: *R*_1_ = 2.1, *R*_2_ = 0.95, *R*_3_ = 2.1 The next *a priori* 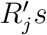 are chosen as: *R*_1_ = 2, *R*_2_ = 0.95, *R*_3_ = 1.9 and we have:

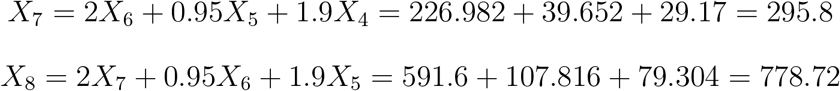 Then, we get the matrices *M*_2_ and 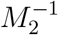

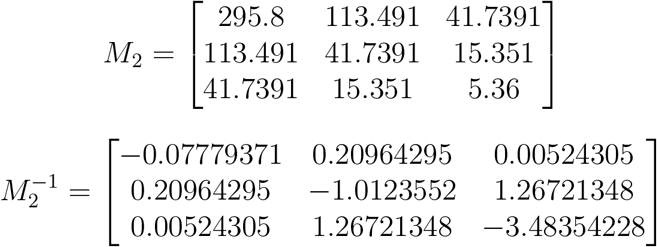 The *a posteriori* 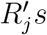 equal: *R*_1_ = 2.0279, *R*_2_ = 7.6158, *R*_3_ = −16.426 The following *a priori* 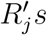 are: *R*_1_ = 2, *R*_2_ = 1.05, *R*_3_ = 1.9 and we have:

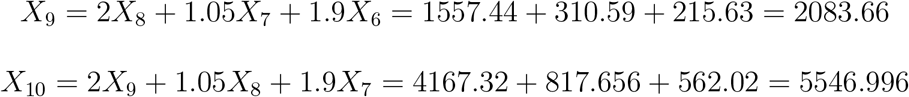 Then, we get the matrices *M*_3_ and 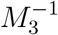

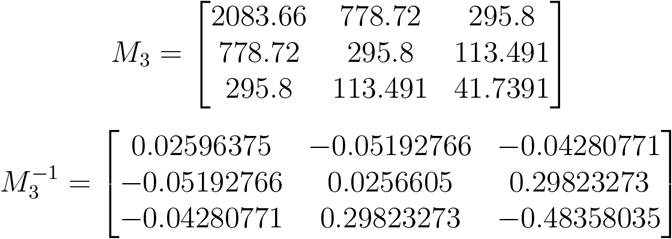 The *a posteriori* 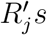 equal: *R*_1_ = 2.486, *R*_2_ = −2.33, *R*_3_ = 7.38769 The next *a priori* 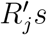 are: *R*_1_ = 1.9, *R*_2_ = 1.05, *R*_3_ = 1.9 and we have:

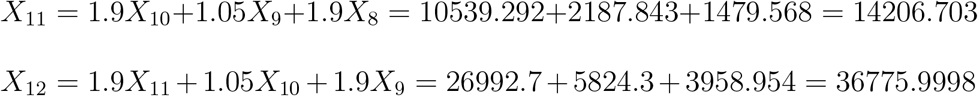 Then, we get the matrices *M*_4_ and 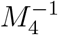

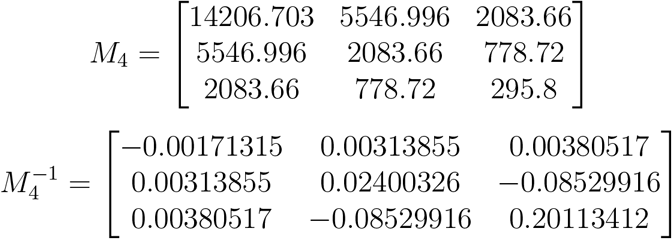 The *a posteriori* 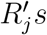 equal: *R*_1_ = 2.69, *R*_2_ = −16.72, *R*_3_ = 43.809
2. **For a=1**, let’s choose the *a priori R*_1_ in [1.0, 3.0], *R*_2_ in [0.5, 1.5] and *R*_3_ in [1, 3] e.g., *R*_1_ = 1, *R*_2_ = 1.355, *R*_3_ = 1.1. Then, transition matrix is equal to:

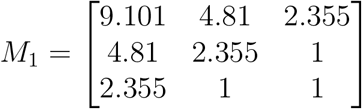

and its inverse is given by:

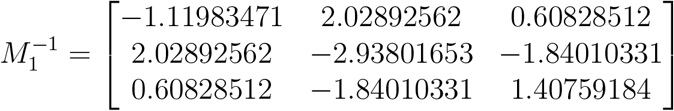

The new cases are: *X*_6_ = 18.209, *X*_5_ = 9.101, *X*_4_ = 4.81, *X*_3_ = 2.355, *X*_2_ = 1, *X*_1_ = 1, and by deconvolution, we get *a posteriori* 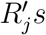 equal: *R*_1_ = 1, *R*_2_ = 1.355, *R*_3_ = 1.1, i.e., the exact *a priori* distribution.

Let’s consider now a new *a priori R*_1_ = 1, *R*_2_ = 1, *R*_3_ = 1. That gives a new matrix *M*_2_, with new *X*_7_ and *X*_8_ calculated from the new *a priori* 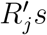, by using the former values of *X*_6_, &, *X*_2_ :

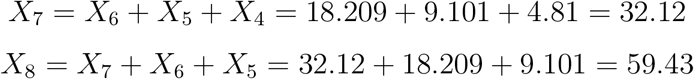

Hence, we get:

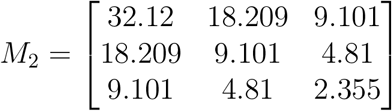

Then, by inverting *M*_2_, we can calculate the *a posteriori* 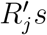:

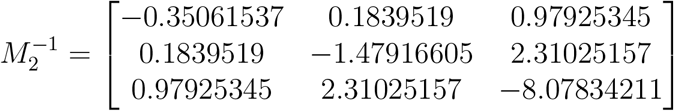

and *a posteriori* 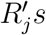 equal: *R*_1_ = 2.90, *R*_2_ = 5.4888, *R*_3_ = −14.696

We continue the process by calculating *X*_9_ and *X*_10_ using new *a priori R*_1_ = 3, *R*_2_ = 0.5, *R*_3_ = 2.9 :

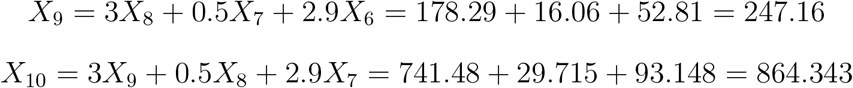

Hence, we get:

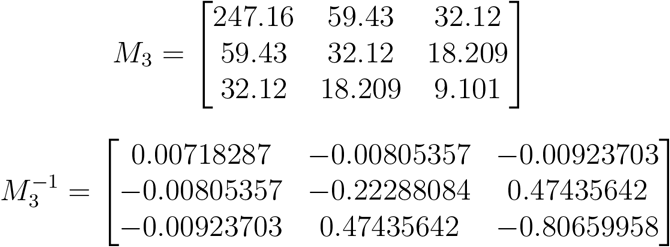

and *a posteriori* 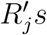 equal: *R*_1_ = 3.66898, *R*_2_ = −33.857, *R*_3_ = 61.32

Let’s choose now a new *a priori R*_1_ = 2.6, *R*_2_ = 0.7, *R*_3_ = 2.6

Then, we have:

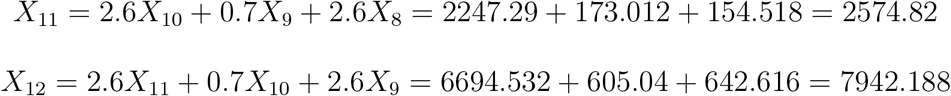

and we get the matrices *M*_4_ and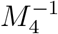 :

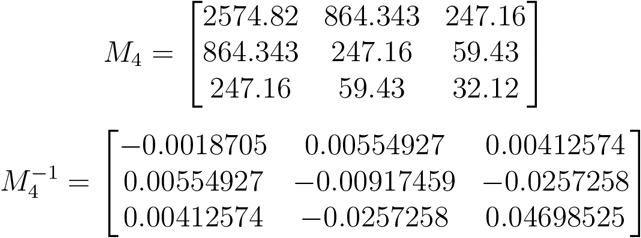

Then, *a posteriori* 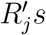 equal: *R*_1_ = 2.99859, *R*_2_ = −1.785, *R*_3_ = 7.139

More precise simulation results are given in Table 1, which summarises computations made for random choices of *a priori* distributions, for *a* = 0.1 and *a* = 1. These simulations show a great sensitivity to the noise, but a qualitative conservation of the down biphasic (D-B) shape of their distribution along the infectivity period of individuals.

**Table 1.** Simulation results obtained for extreme noises *a* = 0.1 and *a* = 1, showing great variations of the deconvoluted *a posteriori* distribution of the daily reproduction numbers 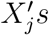 and a qualitative conservation of the down biphasic(D-B) shape of their distribution along the infectivity period. Note: INV means Inverted

| **a** | **a priori $R_j$'s** | **t** | **$X_t$** | **$X_{t+1}$** | **$X_{t+2}$** | **a posteriori $R_j$'s** | **$R_o$** | **D-B** |
| --- | --- | --- | --- | --- | --- | --- | --- | --- |
| 0.1 | 2.1;0.95;2.1 | 4 | 15.35 | 31.74 | 113.5 | 2.1;0.95;2.1 | 5.15 | Yes |
|  | 2;0.95;1.9 | 6 | 113.5 | 295.8 | 778.7 | 2.03;7.6;-16.4 | -6.77 | INV |
|  | 2;1.06;1.9 | 8 | 778.7 | 2083.7 | 5547 | 2.49;-2.33;7.39 | 7.55 | Yes |
|  | 1.9;1.05;1.9 | 10 | 5547 | 14207 | 36776 | 2.69;-16.7;43.8 | 29.8 | Yes |
|  | 1.9;0.95;1.9 | 12 | 36776 | 93910 | 240359 | 2.92;1.68;-6.7 | -2.1 | No |
|  | 1.9;1;1.9 | 14 | 240359 | 622149 | 1605227 | 2.29;-4.83;14.3 | 11.8 | Yes |
|  | 2;1.05;1.9 | 16 | 1605227 | 4331630 | 11561153 | 2.76;27;-70 | -40.2 | INV |
|  | 1.9;1;1.95 | 18 | 11561153 | 29558395 | 76502587 | 2.49;-6.48;17.9 | 13.9 | Yes |
|  | 2;1;2.1 | 20 | 76502587 | 207683519 | 556226772 | 2.67;-7.6;19.7 | 14.8 | Yes |
| 1 | 1;1.355;1.1 | 4 | 4.81 | 9.1 | 18.21 | 1;1.355;1.1 | 3.455 | No |
|  | 1;1;1 | 6 | 18.21 | 32.12 | 59.43 | 2.90;5.49;-14.70 | -6.31 | INV |
|  | 3;0.5;2.9 | 8 | 59.43 | 247.16 | 864.34 | 3.67;-33.9;61.32 | 31.1 | Yes |
|  | 2.6;0.7;2.6 | 10 | 864.34 | 2574.82 | 7942.19 | 3;-1.79;7.14 | 8.35 | Yes |
|  | 2.5;0.75;1.5 | 12 | 7942.2 | 23083.1 | 67526.6 | 3.35;2.54;-11.6 | -5.71 | No |
|  | 2.4;0.8;2.4 | 14 | 67526.6 | 199590 | 588437 | 2.58;-0.5;4.8 | 6.88 | Yes |
|  | 2;1;2 | 16 | 588437 | 1511517 | 4010652 | 2.72;-1.08;3.19 | 4.83 | Yes |
|  | 2.3;1.15;2.3 | 18 | 4010652 | 12316150 | 36415885 | 2.88;-7.9;21.7 | 16.7 | Yes |
|  | 2.8;0.6;2 | 20 | 36415885 | 117375471 | 375133150 | 3.7;4.1;-17 | -9.2 | INV |

### 3.3 Distribution of the daily reproduction numbers R_*j*_’s. The real example of France

The Figure 3 gives the average transmission rate *R*_*o*_ calculated the 25th of October 2020 just before the second lockdown (re-confinement) in France [18]. Because the second wave of the epidemic is still in its exponential phase, it is more convenient i) to consider the distribution of the marginal daily reproduction numbers, and ii) to calculate its entropy and simulate the epidemic dynamics using a Markovian model [2].

By using the daily new infectious cases given by [6], we can calculate *M* ^−1^ for the period from 20th to 25th October 2020, by choosing 3 days for the duration of the infectiousness period and the following raw data for the new infected cases (Figure 3):

Oct 25 : 52010, 45422, 42032, 41622, 26676, 20468 : Oct 20

We have:

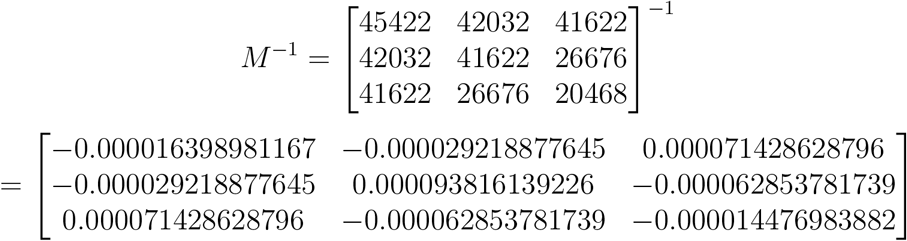

Hence, we can deduce the daily 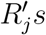 i.e., the vector (*R*_1_, *R*_2_, *R*_3_) :

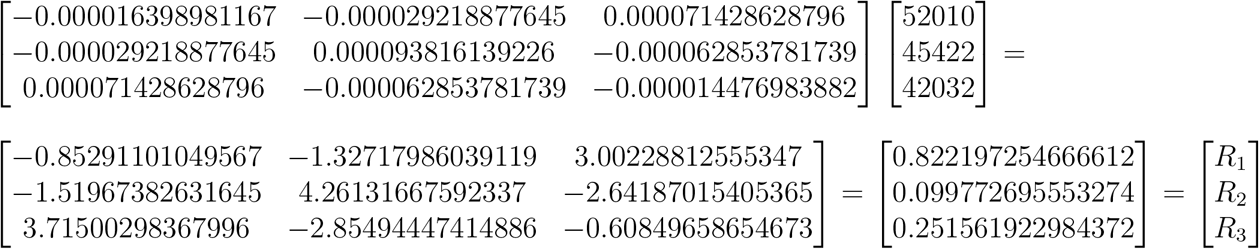

The average transmission rate is equal to *R*_*o*_ ≈ 1, 174, value close to that calculated directly (Figure 3), giving *V* = (0.7, 0.085, 0.215), with a maximal daily reproduction rate the first day of the infectiousness period. The entropy *H* of *V* is equal to: *H* = −∑_*k*=1,*r*_ *V*_*k*_*Log*(*V*_*k*_) = 0.25 + 0.21 + 0.33 = 0.79.

### 3.4 Calculations of the R_*j*_’s for different countries

#### 3.4.1 Chile

By using the daily new infectious cases given by [6], we can calculate *M* ^−1^ for the period from 1st to 12th November 2020, by choosing 6 days for the duration of the infectiousness period and the following 7-day moving average data for the new infected cases (Figure 4):

**Figure 4.**
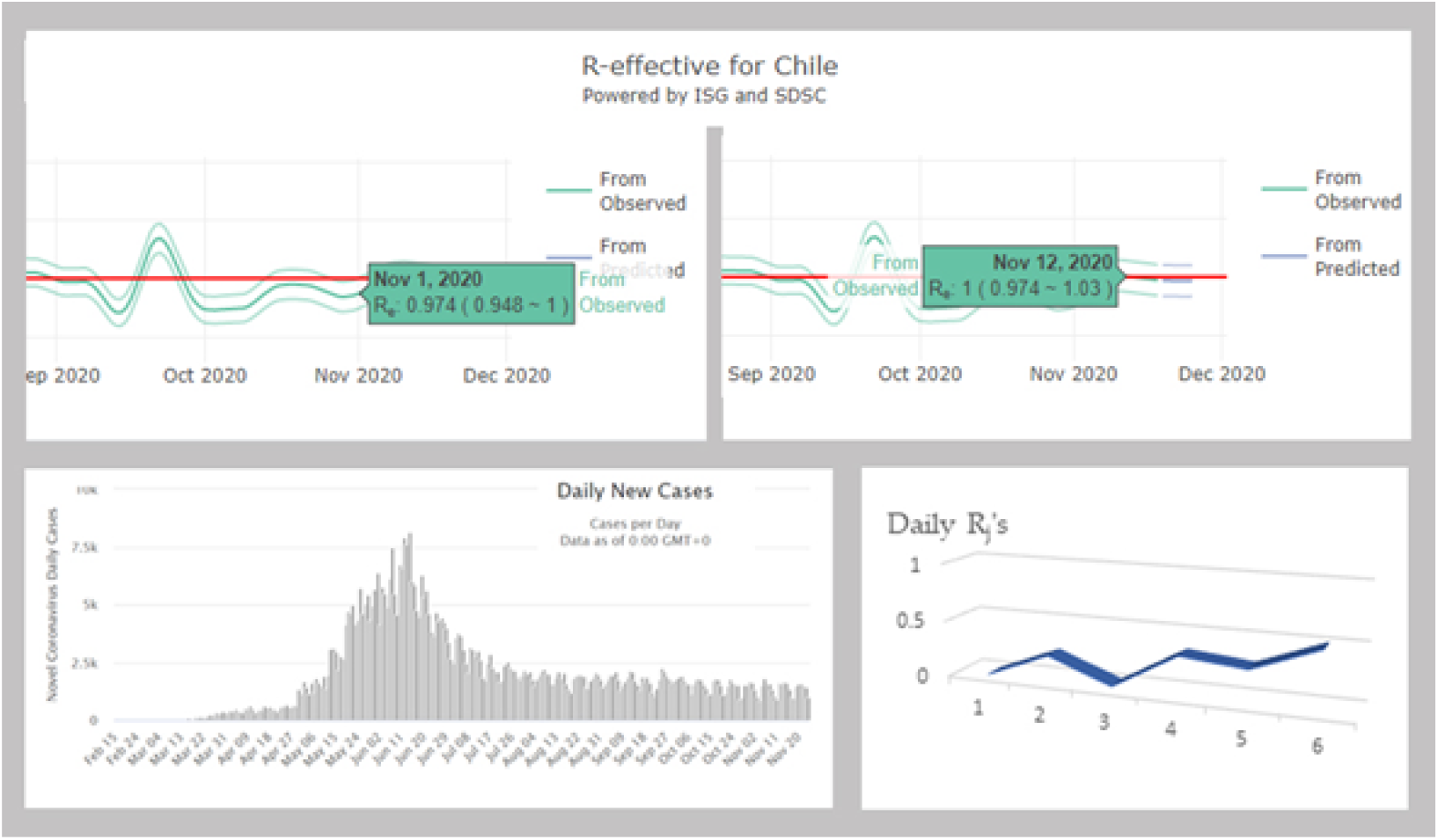
Top: estimation of the average transmission rate *R*_*o*_ for the 1st and the 12th November 2020 [18]. Bottom left: Daily new cases in Chile between November 1 and November 12 [6]. Bottom right: V-shape of the evolution of the daily 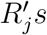 along the infectious 6-day period of an individual.

Nov 12 : 1408, 1394, 1387, 1384, 1385, 1389, 1405, 1362, 1359, 1382, 1370, 1400 : Nov1

We have:

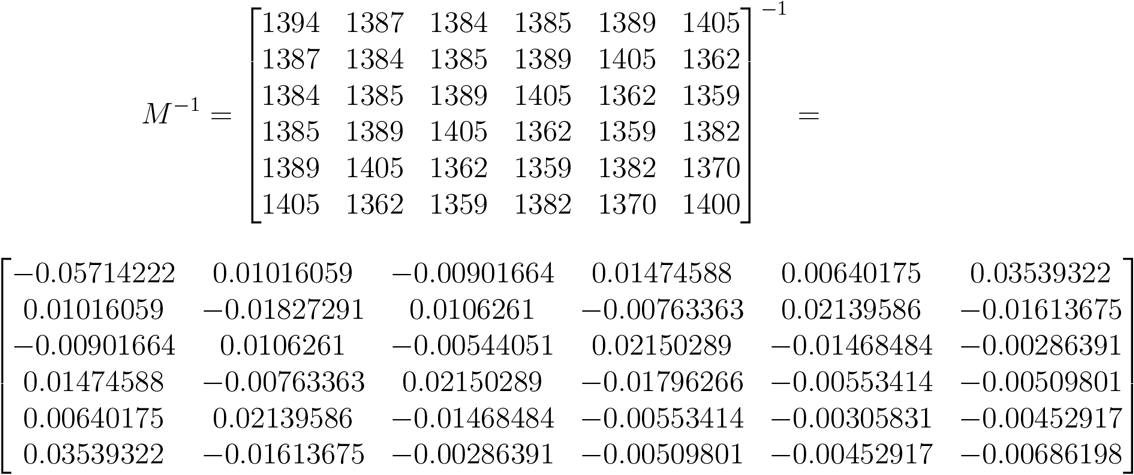

Hence, after deconvolution we have:

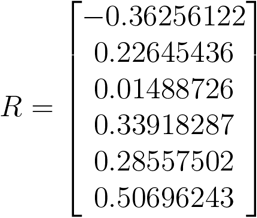

The average transmission rate is equal to *R*_*o*_ ≈ 1, 011, value close to that calculated directly, with a maximal daily reproduction rate the last day of the infectiousness period.

Because of the negativity of *R*_1_, we cannot derive the distribution *V* and therefore calculate its entropy. The quasi-endemic situation in Chile since the end of August, which corresponds to the increase of temperature and drought at this period of the year [15], gives a constancy of new cases of infected and a periodicity of their occurrence corresponding to the length of the infectious period of about 6 days, analogue to the cyclic phenomenon observed in simulated stochastic data of Section 3.2. with a similar M-shaped distribution of the 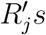.

#### 3.4.2 Russia

By using the daily new infectious cases given by [6], we can calculate *M* ^−1^ for the period from September 30 to October 5, 2020, by choosing 3 days for the duration of the infectiousness period and the following 7-day moving average data for the new infected cases (Figure 5):

**Figure 5.**
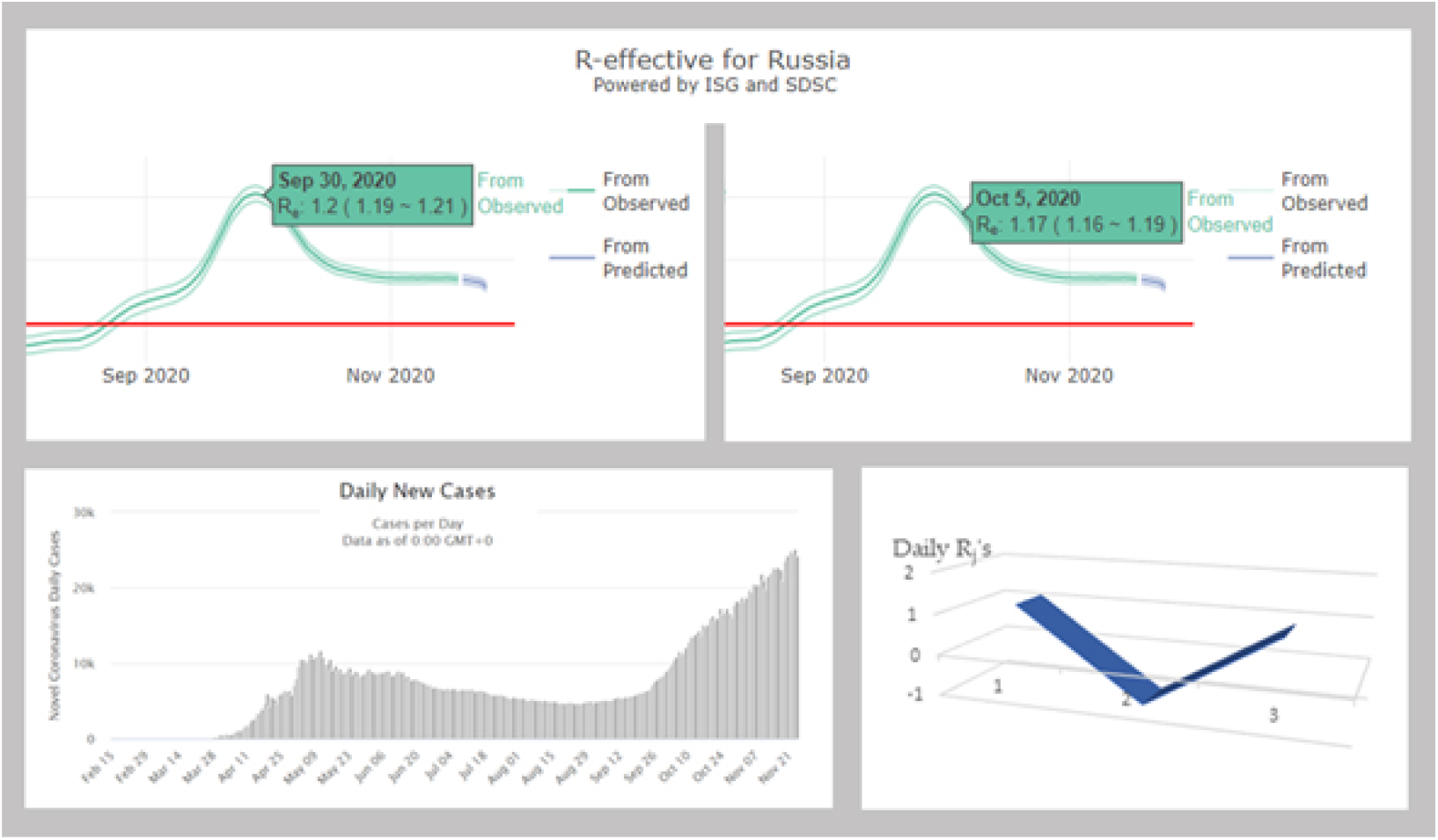
Top: estimation of the average transmission rate *R*_*o*_ for the 30th September and the 5th October 2020 [18]. Bottom left: Daily new cases in Russia between September 30 and October 5 [6]. Bottom right: V-shape of the evolution of the daily 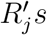 along the infectious 3-day period of an individual.

Oct 05 : 9473, 9081, 8704, 8371, 8056, 7721 : Sept 30

We have:

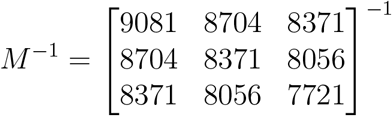

and

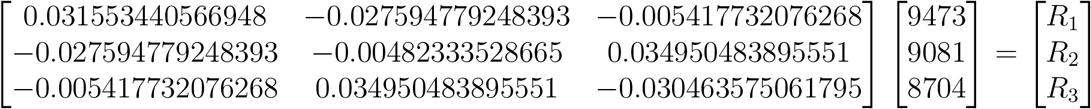

where:

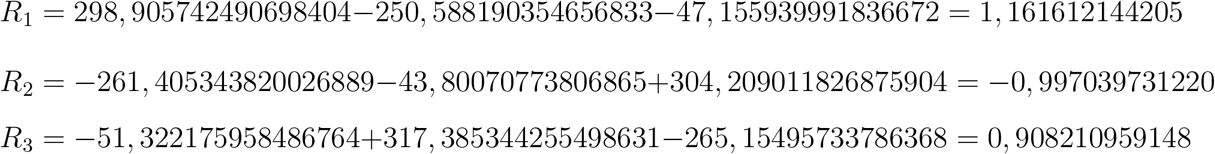

The average transmission rate is equal to *R*_*o*_ ≈ 1, 073, value close to that calculated directly, with a maximal daily reproduction rate the first day of the infectiousness period.

Because of the negativity of *R*_2_, we cannot derive the distribution *V* and therefore calculate its entropy. The period studied corresponds to a local slow increase of new infectious cases at the start of the second wave in Russia, which seems to have a succession of slightly inclined 4-day plateaus followed by a step.

#### 3.4.3 Nigeria

By using the daily new infectious cases given by [6], we can calculate *M* ^−1^ for the period from November 5 to November 10 2020, by choosing 3 days for the duration of the infectiousness period and the following raw data for the new infected cases (Figure 6) :

**Figure 6.**
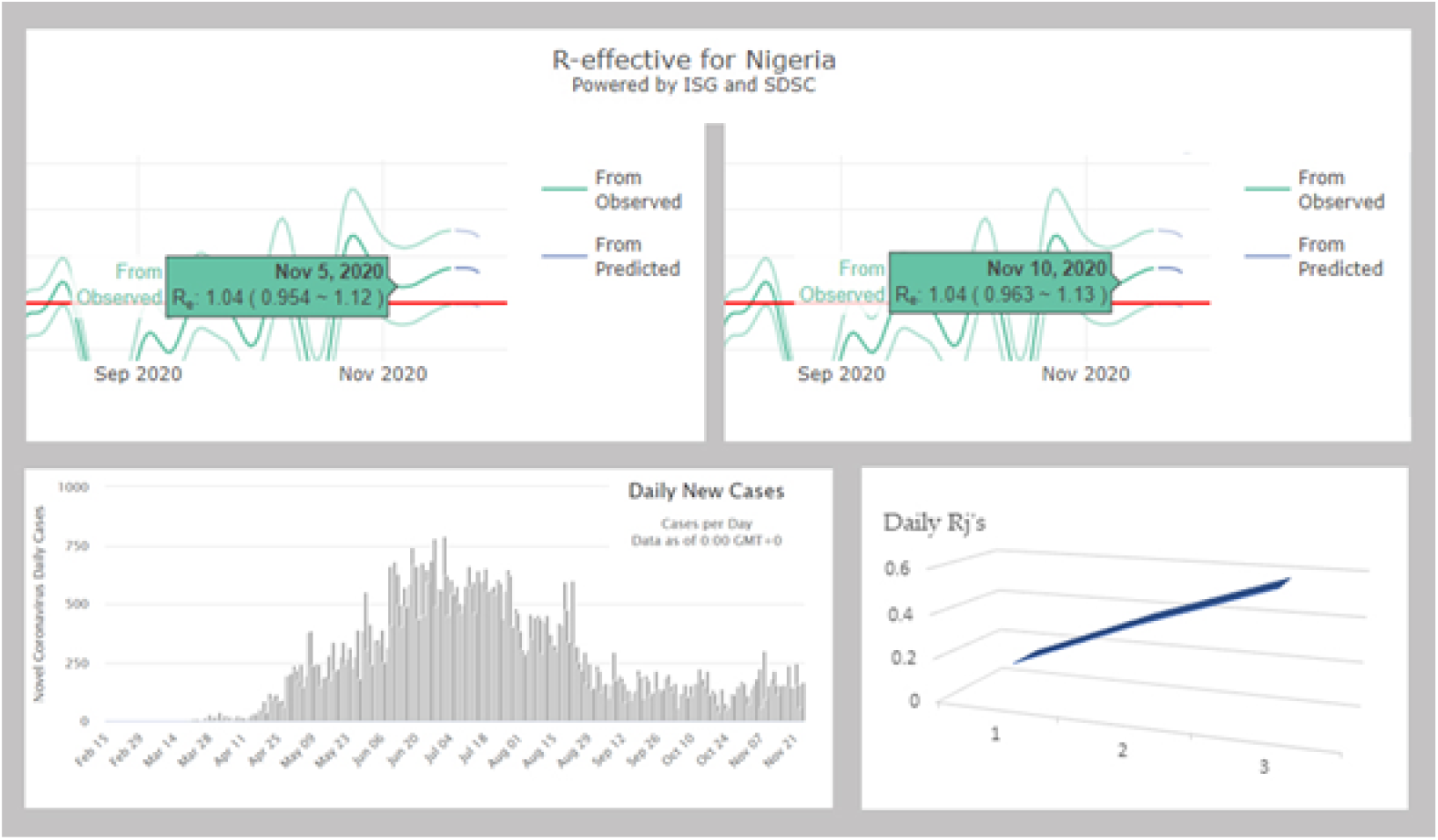
Top: estimation of the average transmission rate *R*_*o*_ for the 5th and the 10th November 2020 [18]. Bottom left: Daily new cases in Nigeria between November 5 and November 10 [6]. Bottom right: increasing evolution of the daily 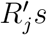 along the infectious 3-day period of an individual.

Nov 10 : 166, 164, 161, 133, 149, 141 : Nov5

We have:

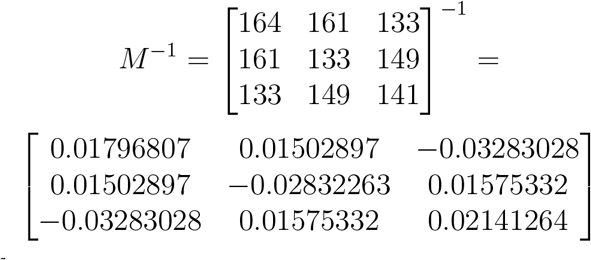

After deconvolution, we get:

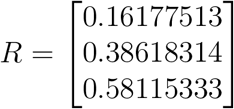

The average transmission rate is equal to *R*_*o*_ ≈ 1.129, value close to that calculated directly, with a maximal daily reproduction rate the last day of the infectiousness period. *V* = (0.143, 0.342, 0.515) and the entropy *H* of *V* is equal to : *H* = −∑_*k*=1,*r*_ *V*_*k*_*Log*(*V*_*k*_) = 0.29 + 0.37 + 0.34 = 1.

### 3.5 Weekly patterns in daily infection cases

Daily new infectious cases are highly affected by weekdays, such that case numbers are lowest at the start of the week, and increase afterwards. This pattern is observed at the global level, as well as at the level of almost each single country or USA state. Hence, in order to estimate biologically meaningful infection rates, clean of weekly patterns due to administrative constraints, analyses have to be restricted to specific periods shorter than a week, or at the rare occasions when patterns escape the administrative constraints on weekly patterns. This weekly phenomenon occurs during exponential increase as well as decrease phases of the pandemic, and during stagnation periods in numbers of daily cases. In addition, the daily new infection case record is discontinuous for many countries/regions, which frequently publish on Monday or Tuesday a cumulative count for that day and the weekend days. For example, Sweden typically publishes over one week only four numbers, the one on Tuesday cumulating cases for Saturday, Sunday and the two first weekdays. Discontinuity in records further limit the availability of data enabling detailed analyses of daily infection rates and can be considered as extreme weekday effects on new case records due to various administrative constraints.

We calculated Pearson correlation coefficients *r* between a running window of daily new case numbers of 20 consecutive days, and a running window of identical duration with different intervals between the two running windows. There is typically a peak with a time-lag of seven days between the two running windows. This could reflect a biological phenomenon of seven infection days. However, examinations of frequency distributions of time-lags for r maxima reveals, besides the median at time-lag 7 days, local maxima for multiples of 7 (14, 21, 28, 35, etc). About 50 percent of all local maxima in *r* involve time-lags that are multiples of seven (seven included). This excludes a biological causation. We tried to control for week days using two methods, and combinations thereof.

For the first method, we calculated z-scores for each weekday, considering the mean number of cases for each weekday, and subtracted that mean from the observed number for a day. This delta was then divided by the standard deviation for the number of cases for that day. This was done for each weekday.

The second method implies data smoothing using a running window of 5 consecutive days, where the mean number of cases calculated across the five days is subtracted from the observed number of cases for the third day. Hence data for a given day are compared to a mean including two previous, and two later days.

We constructed two further datasets, one where the second data-smoothing method is applied to the z-scores from the first method, and the other, where the z-scores from the first method are applied to the data after data-smoothing from the second method (Figures 10 and 11).

**Figure 7.**
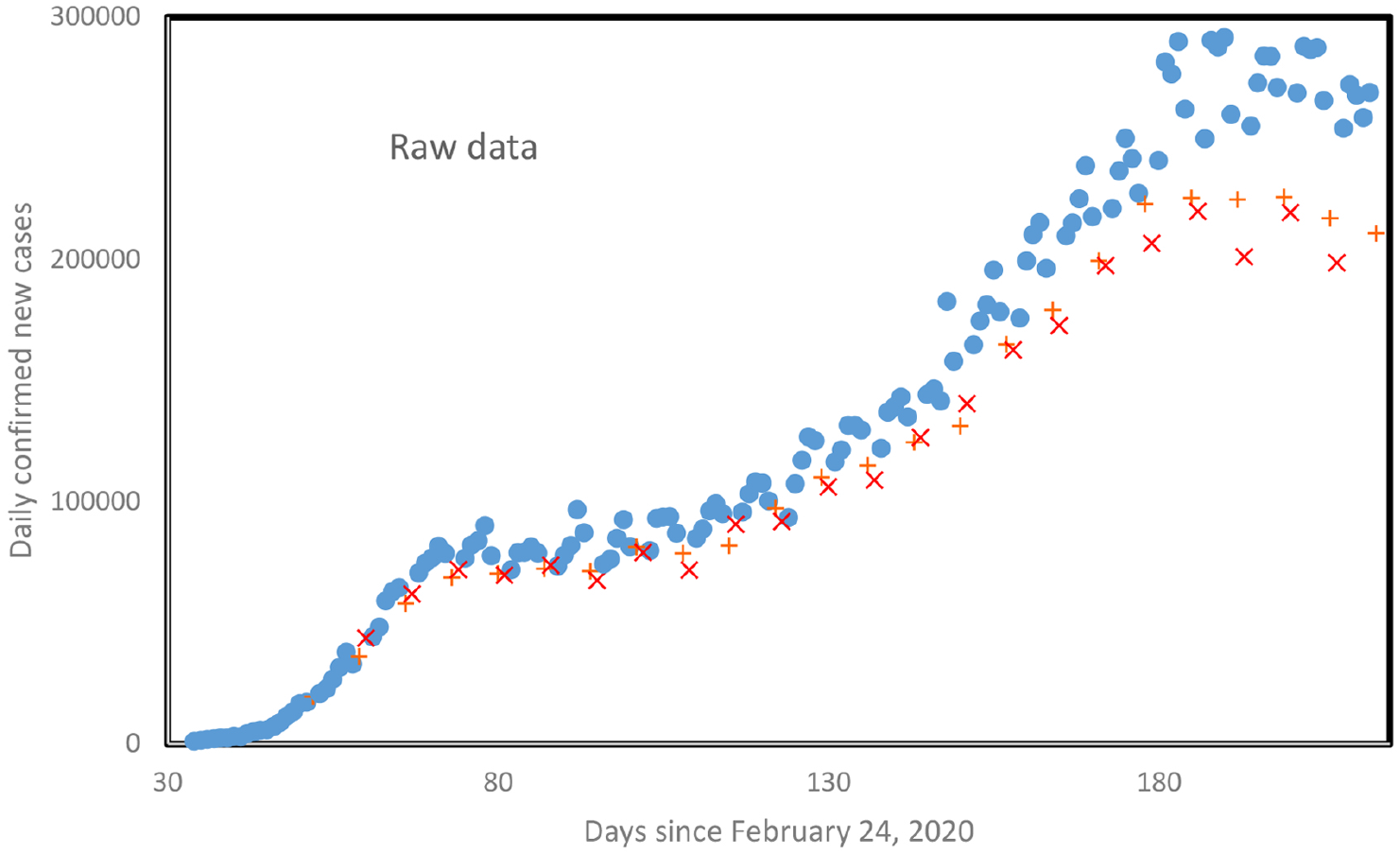
Daily new confirmed cases for the whole world as a function of days since February 24 until August 23.+ indicate Sundays, x indicate Mondays.

**Figure 8.**
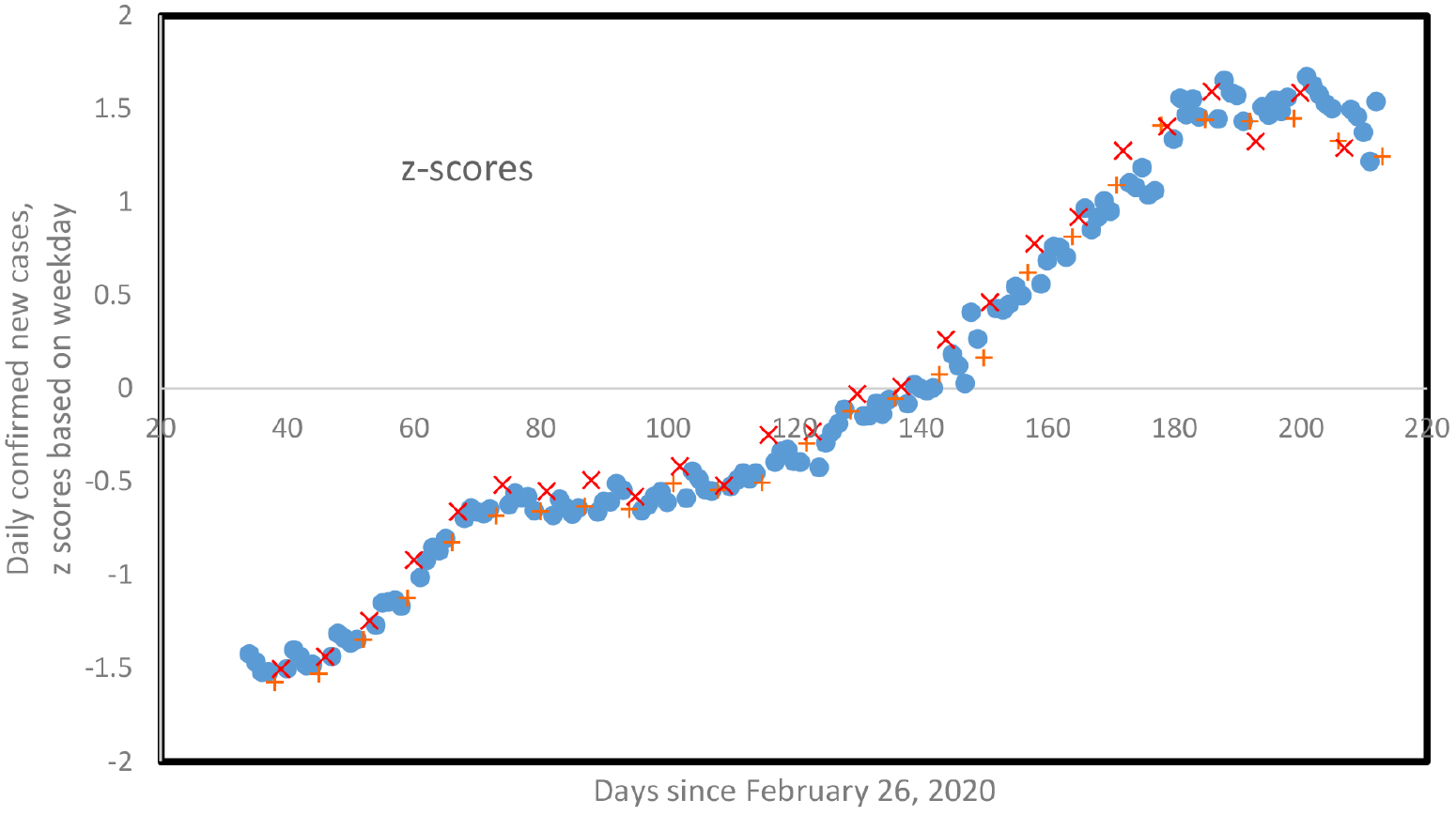
Z-transformed scores of daily new confirmed cases for the whole world, from Figure 7, as a function of days since February 24, until August 23.+ indicate Sundays, x indicate Mondays. Z-transformations are specific to each weekday.

**Figure 9.**
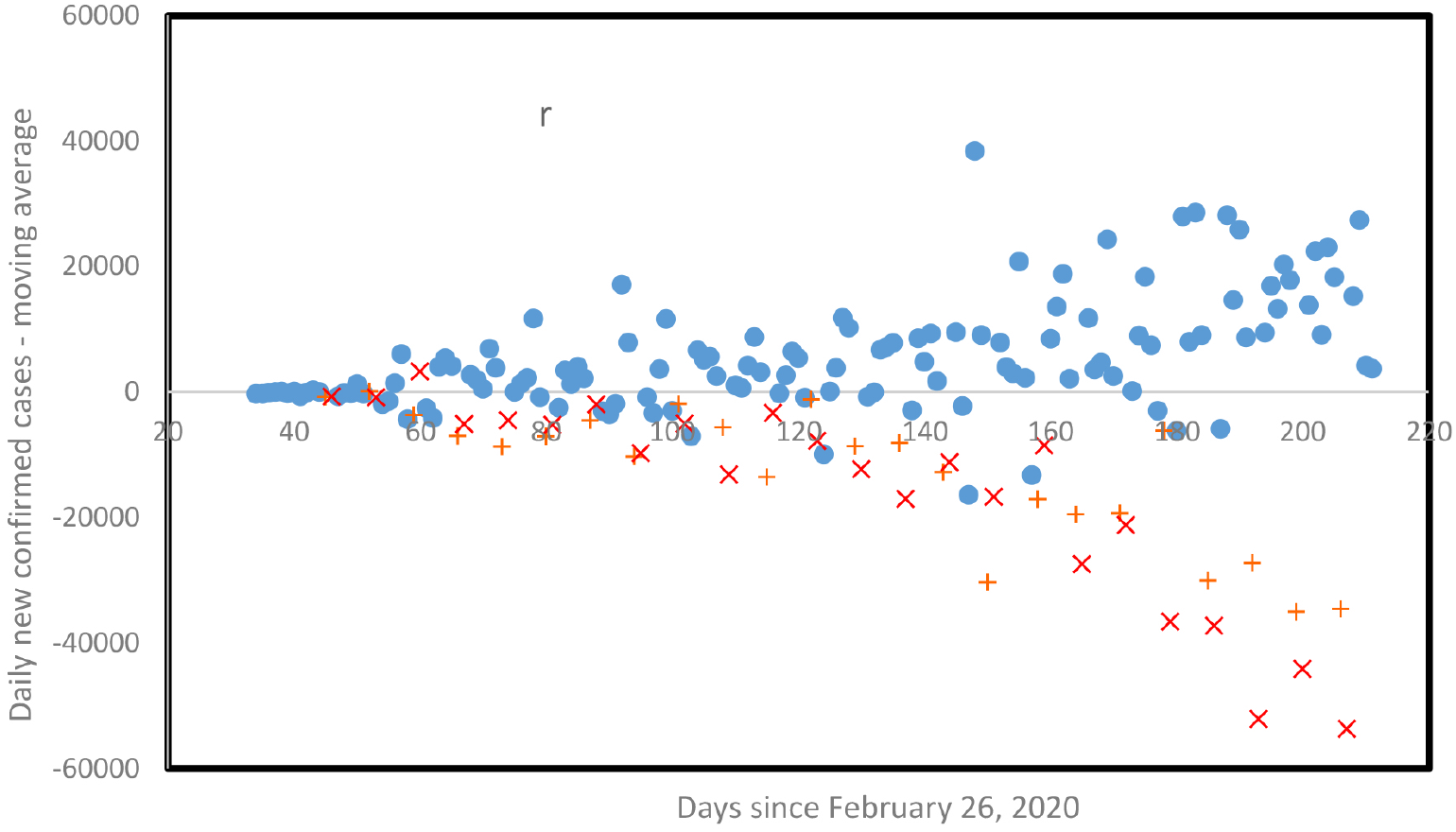
Smoothed daily new confirmed cases for the whole world, from Figure 7, as a function of days since February 24, until August 23. + indicate Sundays, x indicate Mondays. For each specific day *j*, the mean number of new confirmed cases calculated for days *j* − 1, *j* − 2, *j* + 1 and *j* + 2 is subtracted from the number for day *j*.

**Figure 10.**
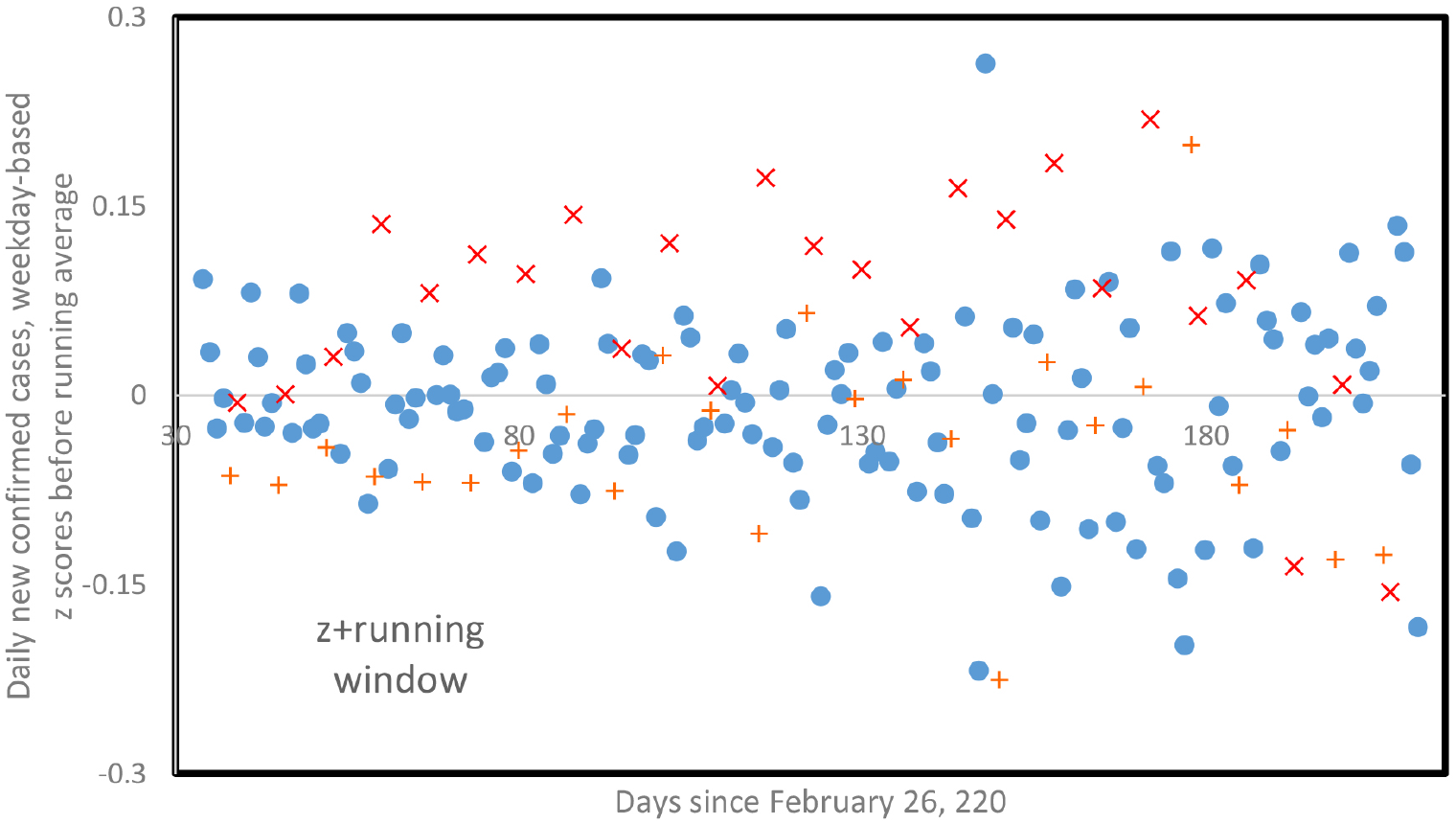
Smoothed daily new confirmed cases for the whole world applied to *Z* scores from Figure 8, as a function of days since February 24, until August 23. + indicate Sundays, x indicate Mondays. For each specific day *j*, the mean number of new confirmed cases calculated for days *j* − 1, *j* − 2, *j* + 1 and *j* + 2 is subtracted from the number for day *j*.

**Figure 11.**
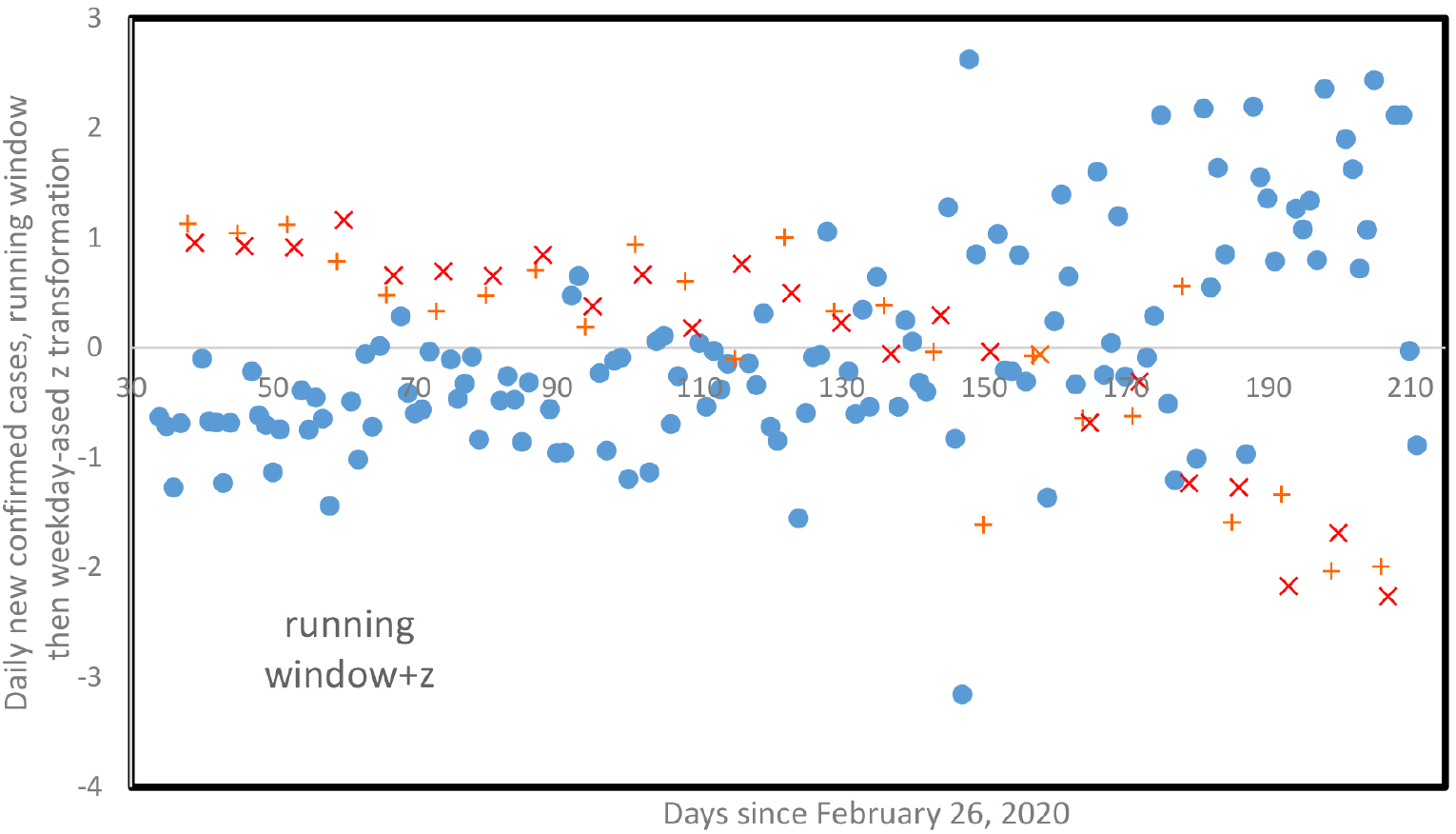
Z-transformed scores of daily new confirmed cases for the whole world, smoothed data from Figure 9, as a function of days since February 24, until August 23.+ indicate Sundays, x indicate Mondays. Z-transformations are specific to each weekday.

These four datasets transformed according to different methods and combinations thereof designed to control for weekday were analysed using the running window method. Despite attempts at controlling for weekday effects, the median time-lag was always seven days across all four transformed datasets, and local maxima in time-lag distributions were multiples of seven. Despite data transformations, about 50 percent of all local maxima were time-lags that are multiples of seven, seven included.

Visual inspection of plots of these transformed data versus time for daily new infection cases from the whole world systematically show systematic local biases in daily new infection cases (after transformations) on Sundays and Mondays, for all four transformed datasets, with Sundays and/or Mondays as local minima and/or local maxima, along which method or combination thereof. Hence, the methods we used failed to neutralise the weekly patterns in daily new cases due to administrative constraints. This issue highly limits the data available for detailed analyses of daily new cases aimed at estimating biologically relevant estimates of infection rates at the level of short temporal scales, but it reinforces the hypotheses done in previous sections on the existence of an heterogeneity of the virulence along the infectiousness period, independent of administrative constraints observed in collecting new daily cases.

## 4 Discussion

Rhodes and Demetrius have pointed out the interest of the distribution *V* of the daily reproduction numbers [5]. In particular, they found that this distribution was generally not uniform, which we have confirmed here by showing many cases with the biphasic form of the virulence observed in respiratory viruses, such as those of influenza. The entropy of the distribution *V* makes it possible to evaluate the intensity of this biphasic character. By taking into account the mortality due to the Covid-19, the discrete dynamics of new cases can be considered as a Leslie dynamics governed by the matrix equation **X**_*j*_ = L**X**_*j*−1_, where **X**_*j*_ is the vector of the new cases living at day *j* and *L* is the Leslie matrix given by:

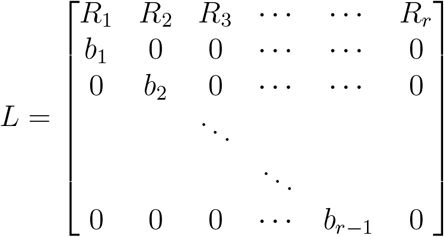

and

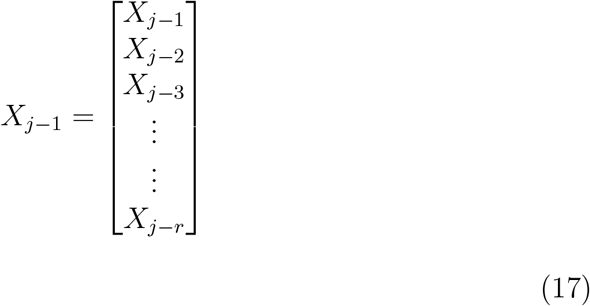

where *b*_*j*_ = 1 − *μ*_*j*_ ≤ 1, ∀*i* = 1, &, *r*, is the survival recovered probability between days *j* and *j* + 1.

The dynamical *X*_*k*_ stability for *L*^2^ distance to the stationary infection age pyramid **P**= *lim*_*j*_**X**_*j*_*/∑* _*k*=*j,j*−*r*+1_ *X*_*k*_ is related to |*λ* − *λ*,|, the modulus of the difference between the dominant and sub-dominant eigenvalues of *L*, namely *λ* = *e*^*R*^ and *λ*, (where *R* is the Malthusian growth rate), where **P** is the left eigenvector of *L* corresponding to *λ*. The dynamical stability for the distance (or symmetrized divergence) of Kullback-Leibler to **P** considered as stationary distribution is related to the population entropy *H* [4,19,20,21,22,23,24], which is defined if *I*_*j*_ = ∏ _*i*=1,*j*−1_ *b*_*i*_ and *p*_*j*_ = *I*_*j*_*R*_*j*_*/λ*^*j*^, as follows:

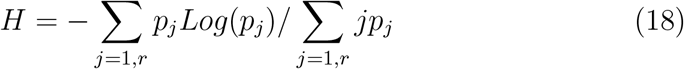

The mathematical characterisation by population entropy defined in (18) of the stochastic stability of the dynamics described by equation (17) has its origin in the theory of large deviations [25,26,27]. This notion of stability pertains to the rate at which the system returns to its steady state conditions after a random exogenous and/or endogenous perturbation and it could be useful to quantify the variations of the distribution of the daily reproduction rates observed for many countries.

## 5 Conclusions and Perspectives

The article has used a discrete approach for describing the dynamics of the Covid-19 outbreak. In the ODE SIR case [28,29,30], marginal 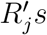 can be estimated in the same way, but by using continuous deconvolution equation. An improvement in this framework could be the introduction of the demographic notion of age class, because the daily reproduction rates distribution is depending on the state of the immune defences both of the infected and susceptible individuals, and on the mode of transmission, this mode being associated in social settings recording age and infection schedule with more secondary cases for young and adult than for elderly at home [31,32]. Then, for each age class *k* a reproduction number *R*_*ok*_ can be calculated by summing the marginal 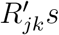, which correspond to this age class *k*, and eventually a global *R*_*o*_ can be estimated from the 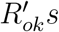, weighted by the proportion of their age class in the whole population. Both global *R*_*o*_ and dedicated to age classes 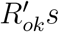 could serve to follow the effects of public health measures as lockdowns and quarantines.

## Data Availability

All data comes from public data bases.

## 6 Author Contributions

Conceptualisation, J.D.; methodology, J.D., H.S.; K.O. and F.T. have performed calculations and Figures; all authors have equally participated to the other steps of the article elaboration.

## 7 Acknowledgements

The authors hereby give their thanks to the International Excellence Fellowship of Karlsruhe Institute of Technology (KIT) funded within the Framework of the University of Excellence Concept “The Research University in the Helmholtz Association I Living the Change”.

## 8 Conflicts of Interest

The authors declare no conflict of interest

## Notes

### Competing Interest Statement

The authors have declared no competing interest.

### Funding Statement

No external funding has been received.

### Author Declarations

All data comes from public data bases, there is no need for IRB nor ethics committee approvals.

